# Frontoparietal functional connectivity underlies top-down attention in mild cognitive impairment

**DOI:** 10.64898/2026.10.30.26365141

**Authors:** Hannah Klink, Kathrin Finke, Fabian Kattlun, Simon J. Schrenk, Marleen Haupt, Stefan Brodoehl, Ha-Yeun Chung, Adriana L. Ruiz-Rizzo

## Abstract

**Background:** Patients with amnestic mild cognitive impairment frequently show attention problems such as reduced top-down control, the ability to ignore task-irrelevant information. It is unclear whether this reduction corresponds with decreased brain connectivity. We aimed to determine whether and how differences in functional connectivity, or the correlated activity between brain areas, explain differences in top-down control in amnestic mild cognitive impairment.

**Methods:** We conducted a case-control study with 26 patients with amnestic mild cognitive impairment (58-91 years; 10 women) and 29 age-matched healthy older adults (60-79 years; 9 women). All patients and control participants underwent neuropsychological assessment to confirm diagnosis and healthy cognitive status, respectively. We used resting-state functional magnetic resonance imaging at 3T to analyse functional connectivity. Top-down control was estimated with a ‘theory of visual attention’-based verbal partial-report task (parameter α; higher α indicates poorer control). We identified visual and attention-relevant functional brain networks and tested for differences between patients and controls while controlling for age and gender.

**Results:** Compared with healthy control participants, patients with amnestic mild cognitive impairment showed lower functional connectivity in the left frontoparietal attention network and in one visual network and worse top-down control α (*t*(37.43) = -2.24, *p* = 0.031). The reduced connectivity in the frontoparietal network, but not the visual network, significantly correlated with top-down control α across all participants (*ρ* = -0.33, *p* = 0.003) and in the patient group (*ρ* = -0.48, *p* = 0.014). Moreover, connectivity in the frontoparietal network mediated the relationship between amnestic mild cognitive impairment and top-down control α.

**Discussion:** These results link lower frontoparietal connectivity to worse attentional control in patients with amnestic mild cognitive impairment. Worse control reflects inefficient attentional selection and increasing distraction by irrelevant information during perceptual processing and encoding into working memory. Overall, this implicates a role for the frontoparietal network in maintaining cognitive control or, more generally, supporting cognitive functioning in older adults with emerging cognitive impairment.

## Introduction

Mild Cognitive Impairment (MCI) constitutes a cognitive impairment beyond what is expected for aging processes in one or more cognitive domains, while functional independence is largely preserved (1). Even though symptoms can remain stable or remit over time, patients with MCI have an increased risk of developing dementia (2,3). Amnestic MCI (aMCI) is characterized by predominant memory impairments (4) and a higher conversion risk to dementia has been reported for multi-domain aMCI in comparison to multi-domain non-aMCI.

Although aMCI is clinically identified by a prominent memory impairment, cognitive deficits are not confined to memory. Studies using established clinical neuropsychological measures have also demonstrated attentional impairments in patients with aMCI (5). Experimental paradigms using response-conflict and oculomotor tasks have further characterized specific difficulties in attentional and inhibitory control in patients with aMCI (6,7). Studies based on the Theory of Visual Attention (TVA) (8) have provided a more process-specific characterisation of impairments in distinct components of visual attention, i.e., in visual-perceptual threshold and visual processing speed (9–11). Of particular relevance to the present study, impaired top-down control of attentional selection has also been reported in aMCI (12). Top-down control enables goal-directed selection by prioritizing task-relevant information over competing distractors and is formalized within the TVA as the relative attentional weighting of distractors compared with targets (8).

Resting-state functional connectivity (rsFC) provides a means of investigating the neural basis of impaired top-down control in aMCI. rsFC based on functional magnetic resonance imaging (fMRI) quantifies the correlation of spontaneous blood oxygenation level dependent (BOLD) signal fluctuations (13,14). Such correlation is meaningful as it reflects dynamic but relatively stable intrinsic functional networks (15). Previous rsFC studies in aMCI (16–18) have mainly focused on alterations of the default mode network (DMN) and their association with memory deficits (19,20). By contrast, considerably less is known about the functional networks underlying specific deficits in attentional selection. Previous findings have implicated frontoparietal areas in impaired top-down control in early stages of Alzheimer’s disease (AD) (21,22). Because top-down selection regulates competition among visual stimuli, altered connectivity within both frontoparietal attention and visual networks may be relevant to impaired top-down control in aMCI. However, direct evidence linking connectivity in these networks to experimentally quantified top-down control in aMCI is missing.

Here, we aimed to identify neural correlates of impaired top-down control in patients with aMCI. In a case-control study, we estimated top-down control using a psychophysical verbal partial-report task based on the theory of visual attention (8) and examined rsFC within attention and visual functional brain networks previously implicated in top-down control (23). These included the dorsal frontoparietal attention network; the lateral frontoparietal attention network; the midcingulo-insular attention network; and visual occipital networks (see Uddin et al. for nomenclature) (15). We hypothesized that, compared with healthy control participants, patients with aMCI would show (i) worse top-down control and (ii) decreased rsFC within these networks, and that (iii) lower rsFC would be associated with poorer top-down control. Finally, to integrate our hypotheses into a model, we tested in a path analysis, whether rsFC mediates top-down control differences between patients with aMCI and healthy control participants.

## Materials and methods

### Participants

Initially, 63 participants took part in our study. Thirty-two patients with a clinical diagnosis of aMCI were recruited from the Memory Centre of the Department of Neurology of Jena University Hospital (Germany) between 2019 and 2024. Thirty-one age- and gender-matched healthy control participants were recruited from the participant pool of our lab between 2021 and 2023. During data curation and preprocessing of magnetic resonance imaging (MRI) data, eight data sets (patients: *n* = 6) were excluded from the analysis due to incomplete resting-state fMRI (*n* = 3) and missing partial-report task data (*n* = 3), extensive head motion (mean framewise displacement exceeding 0.5 mm) during scanning (*n* = 1) or failure to perform the partial-report task (*n* = 1). The final sample consisted of 55 participants (age = 69.9 ± 7.06, 19 females; Table 1), 26 of whom were patients.

**Table 1.**
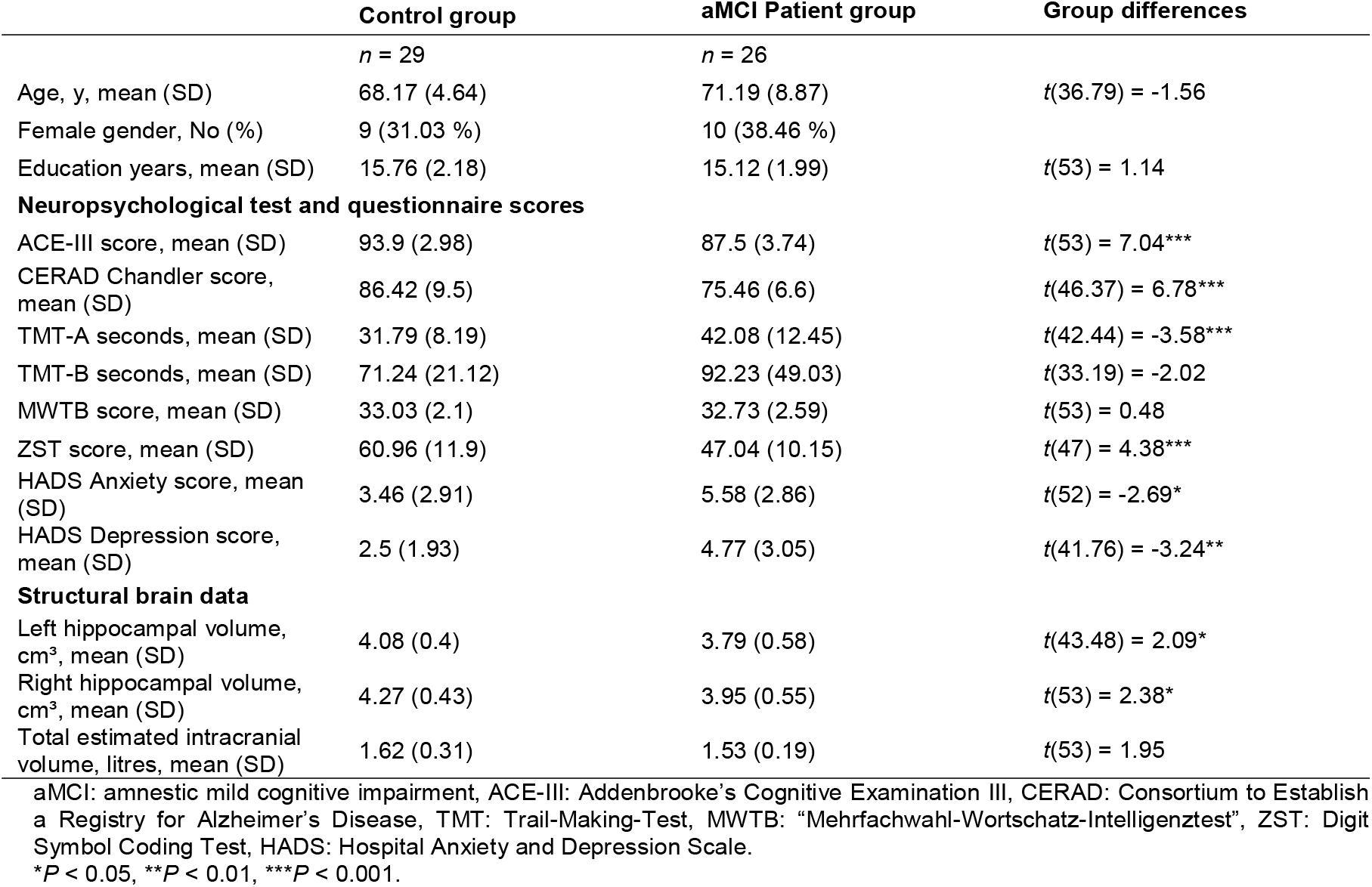
Baseline characteristics by group.

Cerebrospinal fluid (CSF) biomarker and clinical MRI data were available for 20 (76.9 %) and 23 (88.5 %) patients, respectively. Out of the 20 patients with both CSF and MRI data available, six patients showed an AD-consistent amyloid-positive CSF profile, whereas 15 showed clinical MR signs of neurodegeneration (see Additional file 1). All study participants had normal or corrected-to-normal vision and were compensated for their participation. Written informed consent was obtained from all participants. The study was reviewed and approved by the ethics committee of Jena University Hospital, and it was conducted according to the ethical principles outlined in the Declaration of Helsinki II.

### Procedure

#### Neuropsychological assessment

All patients underwent a standard diagnostic routine with expert neurologists and neuropsychologists from the Memory Centre of the Department of Neurology of Jena University Hospital. The neuropsychological assessment included the Addenbrooke’s Cognitive Examination III (ACE-III) (24) and the Consortium to Establish a Registry for Alzheimer’s Disease (CERAD) (25) to assess global cognition with a focus on memory; the Digit Symbol Substitution Test (ZST) (26) and the Trail-Making-Test A and B (TMT-A and TMT-B) (27) to test processing speed and task switching, respectively; the Stroop task (FWIT) (28) to assess executive function; and the “Mehrfachwahl-Wortschatz-Intelligenztest” version B (MWT-B) (29) to test crystallized intelligence. Additional measures included a patient interview (life and medical history, current complaints), sleep and psychiatric questionnaires (Pittsburgh Sleep Quality Index, PSQI; (30); Hospital Anxiety and Depression Scale, HADS-D; (31); Geriatric Depression Scale, GDS (32)), informant questionnaires (Behavioural Assessment of the Dysexecutive Syndrome Questionnaire, BADS-DEX; (33); Informant Questionnaire on Cognitive Decline for the Elderly 7 item version, IQCODE; (34); Bayer Activities of Daily Living Scale, BADL (35)), and the Clinical Dementia Rating (CDR) (36). Collection of CSF was optional and thus performed on 20 patients. Key criteria for aMCI diagnosis included objective cognitive impairment, defined as performance at least 1.5 standard deviations below age-adjusted normative values in at least one neuropsychological measure, with memory being affected. In addition, patients had largely preserved activities of daily living and a CDR score of 0.5. Alternative neurological or psychiatric conditions that could sufficiently explain the cognitive impairment were excluded based on the clinical assessment. Control group participants were assessed with similar neuropsychological tests as those used for patients to ensure that their cognitive status was unimpaired. The exact tests used, however, depended on the respective study protocol (Fit4Brain (37) vs. ABC-MCI (38)). Control participants in both studies completed the ACE-III, TMT and HADS-D questionnaire. Additionally, controls in the Fit4Brain study were assessed with the FWIT, the Verbal Learning Memory Test (VLMT)(39) and the Rey Osterrieth Complex Figure Test (ROCFT; visuospatial processing and memory)(40). Those control participants from the Fit4Brain study assessed for the current study additionally completed the CERAD, ZST and MWTB. Control participants in ABC-MCI regularly completed the CERAD, ZST, and MWTB.

#### Measurement and estimation of top-down control α

We used a verbal partial-report task based on the Theory of Visual Attention (TVA, Fig. 1) (8) similar to previous studies of our research group (37,41). Participants sat in a dimly lit room in front of a computer screen (100-Hz refresh rate) at a fixed distance of 55 cm with their head on a chin rest. During each trial, they were presented with up to two randomly chosen letters from a prefixed set (“ABCDEFGHJKLMNOPRSTUVWXZ”). The letters appeared for a short individual exposure duration (average exposure duration 127.27 ± 38.37 ms), which was determined in a pre-test of two runs with 24 trials each. The task was to name only the red letters (targets) and ignore the blue letters (distractors) (see Fig. 1). Each trial presented either one target, two targets, or one target and one distractor. The letters were arranged in an imaginary square and appeared at the corners. In trials with two letters, both were always arranged next to or above each other, never diagonally. Each trial was preceded by a fixation point, which participants were instructed to fixate upon until letters were presented. Letters were succeeded by blue-red masks to avoid afterimages, followed by a black screen to prompt the participant’s response, who could name letters in arbitrary order and without time pressure. The next trial started after the experimenter had logged the participant’s response. After 13 example trials in the beginning and the two pre-test blocks to determine the exposure duration for each participant, the main task consisted of six test blocks of 48 trials each, resulting in 288 trials in total. Participants were allowed short breaks in between the blocks. The overall task took on average 30 min to complete, including instructions, pre-test blocks and breaks.

**Figure 1.**
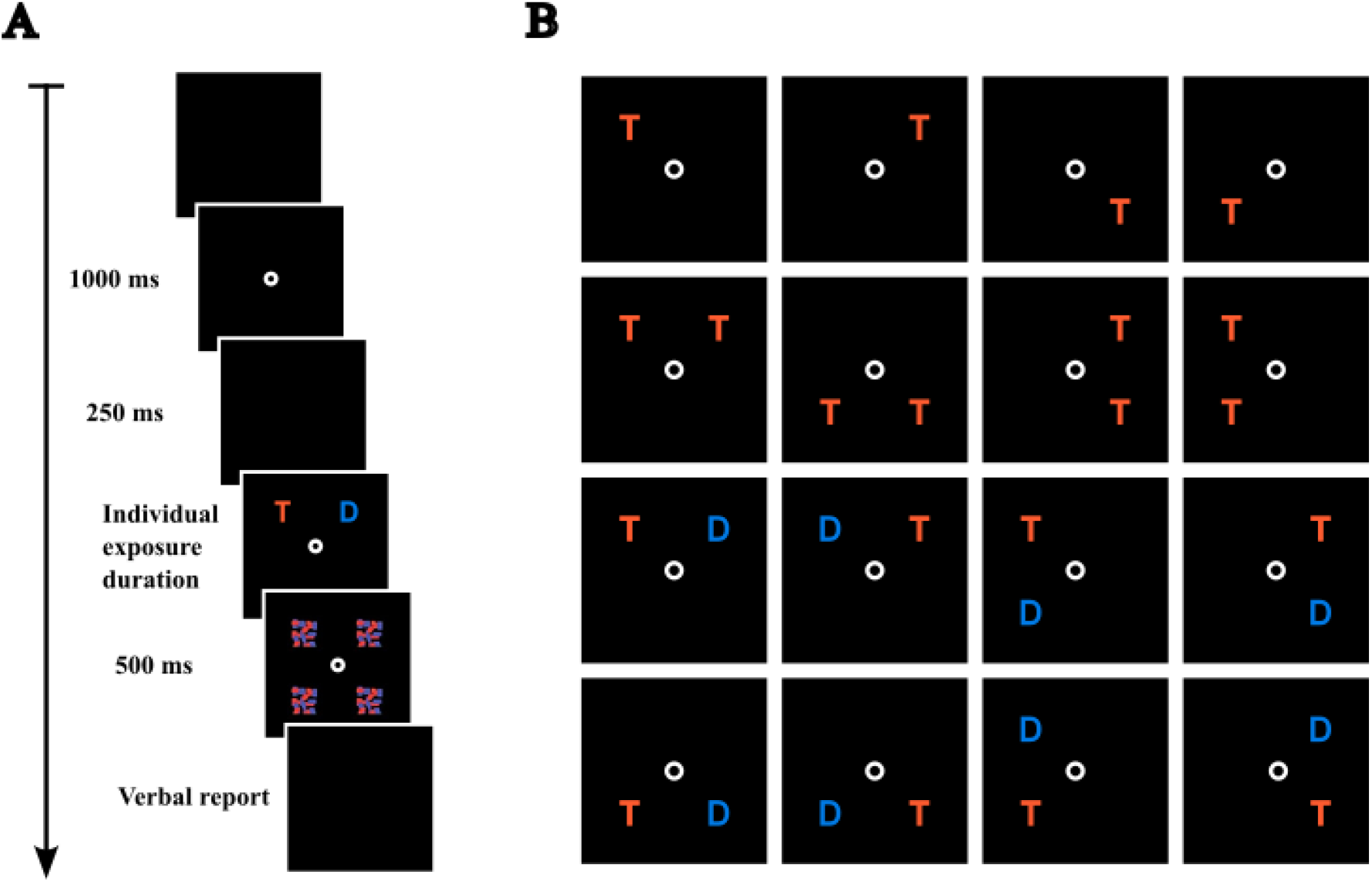
Schematic of the TVA-based partial report task. (**A**) After the presentation of the fixation point for 1000 ms and a blank screen of 250 ms the letters are displayed with an individual exposure duration determined in a pre-test (average exposure duration 127.27 ms ± 38.37). The items are then masked for 500 ms, after which a blank screen prompts the participant to verbally report the target (red letter) only. (**B**) There is a total of 16 different item display types, depending on placements and target-distractor configurations (https://doi.org/10.6084/m9.figshare.33403348). **Figure 1. Alt text.** Figure labelled A and B. A is depicting a single example trial via a sequence of six black boxes as computer screens. The sequence depicts a blank screen, the fixation point, another blank screen, the letters, then the masks and last, another blank screen. B depicts the possible display types of letters via 16 black boxes. Either one target letter, two target letters or one target letter and one distractor letter are arranged around the fixation point in 16 possible combinations resulting from placing the letters either horizontally below or above the fixation point or vertically left or right to the fixation point.

Using TVA functions (42), the model was fitted individually to the observed partial-report task data using MATLAB R2017a (The MathWorks, Inc.). Top-down control was estimated by the attentional selectivity parameter α, which represents the ratio between the attentional weight given to distractors over the attentional weight given to targets (*W*_distractors_/*W*_targets_). Targets receiving more attentional weight than distractors would indicate selective processing and result in α < 1. Unselective processing would result in attentional weights being equally distributed between targets and distractors, α = 1. Thus, values of α closer to 1 represent *worse* top-down control (less selectivity), while values closer to 0 represent *better* top-down control. Values of α > 1 would indicate a prioritisation of processing task-irrelevant distractors.

### Magnetic resonance imaging (MRI) data

#### MRI data acquisition

Brain imaging was acquired on a 3.0 T Siemens MAGNETOM Trio MR scanner (Siemens Healthcare, Erlangen, Germany). During data collection, the acquisition parameters of the T1-weighted anatomical magnetization-prepared rapid gradient echo (MP-RAGE) sequence were updated after 17 patients had been measured. All subsequent patients (n = 15) as well as all control participants (n = 31) were measured with the updated sequence. The earlier and updated sequence parameters were, respectively, repetition time, TR = 2300/2400 ms, echo time, TE = 2.07/2.22 ms, voxel size = 1.0/0.8 mm^3^ isotropic, flip angle = 9°/8°, field-of-view, FOV = 256/256 mm, matrix size = 256/320 × 256/320, total acquisition time = 6 min 38 s.

Resting-state functional images were acquired on the same scanner using identical parameters throughout the study. Functional MRI data were obtained with a T2*-weighted Echo-Planar-Imaging (EPI) sequence acquiring 250 volumes during resting state (TR = 1950 ms, TE = 34 ms, voxel size = 1.4 × 1.4 × 1.4 mm^3^, flip angle = 90°, FOV = 224 mm, matrix size = 160 × 160, phase encoding direction = anterior to posterior, total acquisition time = 8 min 43 s, multiband factor = 4). Each functional volume included 100 trans-axial slices covering the entire brain (cerebellum included). We instructed all participants to close their eyes, lie still, and stay awake during the functional sequence. The signal-to-noise ratio of the functional images did not differ between the two different anatomical acquisition settings (*t*(56) = 0.05, *p* = 0.959).

#### Functional and structural MRI data preprocessing

All brain images were first brain-extracted for further processing with the *bet* command of FSL (FSL version 6.0.5) (43). Each anatomical image was first cropped and bias-field corrected using the *fsl_anat* command. Then, each participant’s anatomical image was brain-extracted by individually adjusting the fractional intensity threshold (*f*) and the threshold gradient (*g*) parameters. Individual adjustment was necessary due to widespread aging and pathological brain changes in the data (e.g. general atrophy), which precluded an adequate brain extraction with the default parameters. Resting-state fMRI data were then brain-extracted using the parameters from the anatomical image. Further preprocessing of resting-state fMRI data was done with the FEAT toolbox of FSL (FEAT v6.00). We discarded the initial five volumes to account for signal stabilisation. Data were motion-corrected with 3 rotations and 3 translations in MCFLIRT (44) and slice-time corrected. Functional images were registered to the individual anatomical images using Boundary-Based Registration (BBR) and normalized to MNI (Montreal Neurological Institute) space with a 2-mm isotropic voxel size using linear transformations with 12 DOF. We used a 6-mm full-width-at-half-maximum Gaussian kernel for spatial smoothing. As resting-state fMRI data are particularly susceptible to motion artifacts, we applied independent component analysis (ICA)-based automatic removal of motion artifacts (ICA-AROMA) (45,46) to the data. ICA-AROMA decomposes the data into independent components and compares those against a set of a priori defined head motion artifacts. This classification is based on high-frequency content, correlation with realignment parameters, edge fraction and CSF fraction. Components labelled as motion artifacts were regressed out non-aggressively and the cleaned data were used for further processing. After applying ICA-AROMA, we band-pass filtered the data using a 100-s cut-off (0.01 Hz) to reduce the effect of low-frequency drifts. Anatomical images were pre-processed in FreeSurfer (version 7.4.1,) (47,48) using the *recon-all* command.

#### Functional connectivity and structural analysis

rsFC analysis was carried out in FSL MELODIC (version 3.15) (49,50). We used group-ICA to extract spatiotemporal signal components representing intrinsic brain networks on a group level, reducing data to 75 dimensions following Allen et al. (51). The 75 group-level independent components (ICs) were then regressed back onto the individual participants in a dual regression analysis (*dual_regression* v0.6) in two stages (temporal and spatial), resulting in individual spatial maps with associated time courses. To identify components representing established intrinsic brain networks, we visually inspected and spatially matched the group-ICA components to the ICA-based resting-state network templates of Allen et al. (51) using the *fslcc* command. All components above and equal to a correlation coefficient of 0.3 were identified as brain networks. For the group analyses, we focused on attention and visual networks only (23). Next, to confirm our network selection, we used the recently released Network Correspondence Toolbox (NCT) (52). The output from FreeSurfer *recon-all* was used for the acquisition of volumetric information about the left and right hippocampi and total intracranial volume.

#### Cluster analysis

We used the FSL *randomise* command with the default setting of 5000 permutations and controlled for age and gender during a voxel-wise analysis within each network of interest to test for rsFC differences between patients and controls. We applied threshold-free cluster enhancement (TFCE) (53) to identify clusters within the attention and visual networks, where patients and control participants significantly differed in their rsFC values and further applied multiple comparisons correction through a false discovery rate of 0.05 to account for the several individual networks tested. Participants’ rsFC values were extracted from all significant clusters. Clusters’ anatomical locations were determined according to the Harvard-Oxford Cortical Structural Atlas available in *fsleyes* (version 1.19.0). The identified clusters were then correlated with top-down control α values (see *Statistical Analyses*).

### Statistical analyses

We used unpaired Student’s *t*-tests to investigate group differences between patients and controls as well as patient differences regarding biomarkers in all variables of interest. In case of unequal variances, we used Welch’s *t*-test instead. Variables of interest included neuropsychological test results, hippocampal volume, top-down control α as the primary outcome variable, and demographic variables. Patients or controls with missing data in key variables (top-down control α, functional connectivity) were excluded from analyses during data curation. Out of the 55 included study participants, only four controls had missing data on either one or two neuropsychological tests. As their cognitive status could still be reliably ascertained, all four controls were left in the analysis.

To test our main hypothesis of an association between functional brain connectivity and top-down control α, we used correlation analyses. Those were calculated with Spearman’s *ρ* coefficient due to violation of Pearson correlation assumptions for some variables or groups. 95 % confidence intervals (CI) were estimated using the Fieller, Hartley and Pearson method (54). Correlation analyses were carried out for the whole sample and both groups separately.

In case the correlation results differed between groups in terms of significance, we computed partial correlations using Group as a control variable to compare those differences statistically.

As control analyses, we computed partial correlations controlling for age, total years of education and depressive symptoms (HADS-D score) to account for potential confounding by these variables. The 95 % confidence intervals were estimated in a bootstrapping procedure with 5000 iterations. Additionally, we tested, *post hoc*, potential brain hemispheric asymmetries in the rsFC results involving group differences and correlations.

Finally, in an exploratory mediation analysis, we tested whether rsFC in the rsFC cluster(s) showing patient-control difference(s) statistically mediated the association between group and top-down control α.

We further tested whether patients differed in variables of interest according to AD biomarkers and evidence of neurodegeneration (CSF and MRI; see Additional file 2). The data were taken from clinical records and categorized into three groups (‘pathology present,’ ‘pathology not present’ and ‘unclear’) based on medical reports. The label ‘unclear’ was given to patients with elevated tau levels indicating tau pathology without Amyloid β burden. Patients with unclear AD pathology were excluded from the analysis.

For all analyses, results were deemed significant at an alpha level of 0.05 and a 95% CI based on bootstrapping was used to assess the statistical significance of mediation parameters. Statistical analysis was carried out with SPSS (version 2025.05.1 Build 513, IBM, Chicago, IL) and mediation analyses were carried out with the PROCESS macro for SPSS (version 5.0), a regression-based tool for estimating direct and indirect effects in a mediation model (55). Boxplots and correlation figures were created with R (version 4.5.1, R Core Team) and RStudio (version 2025.05.1, Posit PBC, Boston, MS).

## Results

### Demographic and clinical variables

Means and standard deviations of all test results for both groups are shown in Table 1. There were no significant age differences between groups (*t*(36.79) = -1.56, *p* = 0.128). Total years of education also did not differ between groups (*t*(53) = 1.14, *p* = 0.260). As expected, patients with aMCI showed worse cognitive performance than healthy control participants in most neuropsychological tests, including the ACE-III (*t*(53) = 7.04, *p* < 0.001) and CERAD (*t*(46.37) = 6.78, *p* < 0.001), as well as higher anxiety (*t*(52) = -2.67, *p* = 0.010) and depressive symptoms (*t*(41.76) = -3.24, *p* = .002). Patients with aMCI showed smaller hippocampal volumes in both hemispheres (left: *t*(43.48) = 2.09, *p* = .043; right: *t*(53) = 2.41, *p* = .010) than healthy controls, despite similar total estimated intracranial volume (*t*(53) = 1.95, *p* = .057). No group differences were found for the TMT-B (*t*(33.19) = -2.01, *p* = 0.051) and MWTB (*t*(53) = 0.48, *p* = 0.633).

### Top-down control α

Patients with aMCI exhibited worse top-down control α (*t*(37.43) = -2.24, *p* = .031, Fig. 2, Table 2). Top-down control α did not significantly correlate with demographic variables (age, education), neuropsychological test results (ACE-III, CERAD), anxiety/depressive symptoms, or hippocampal volume in the total sample or in any group (all *p*-values ≥ 0.083; see Additional file 3).

**Figure 2.**
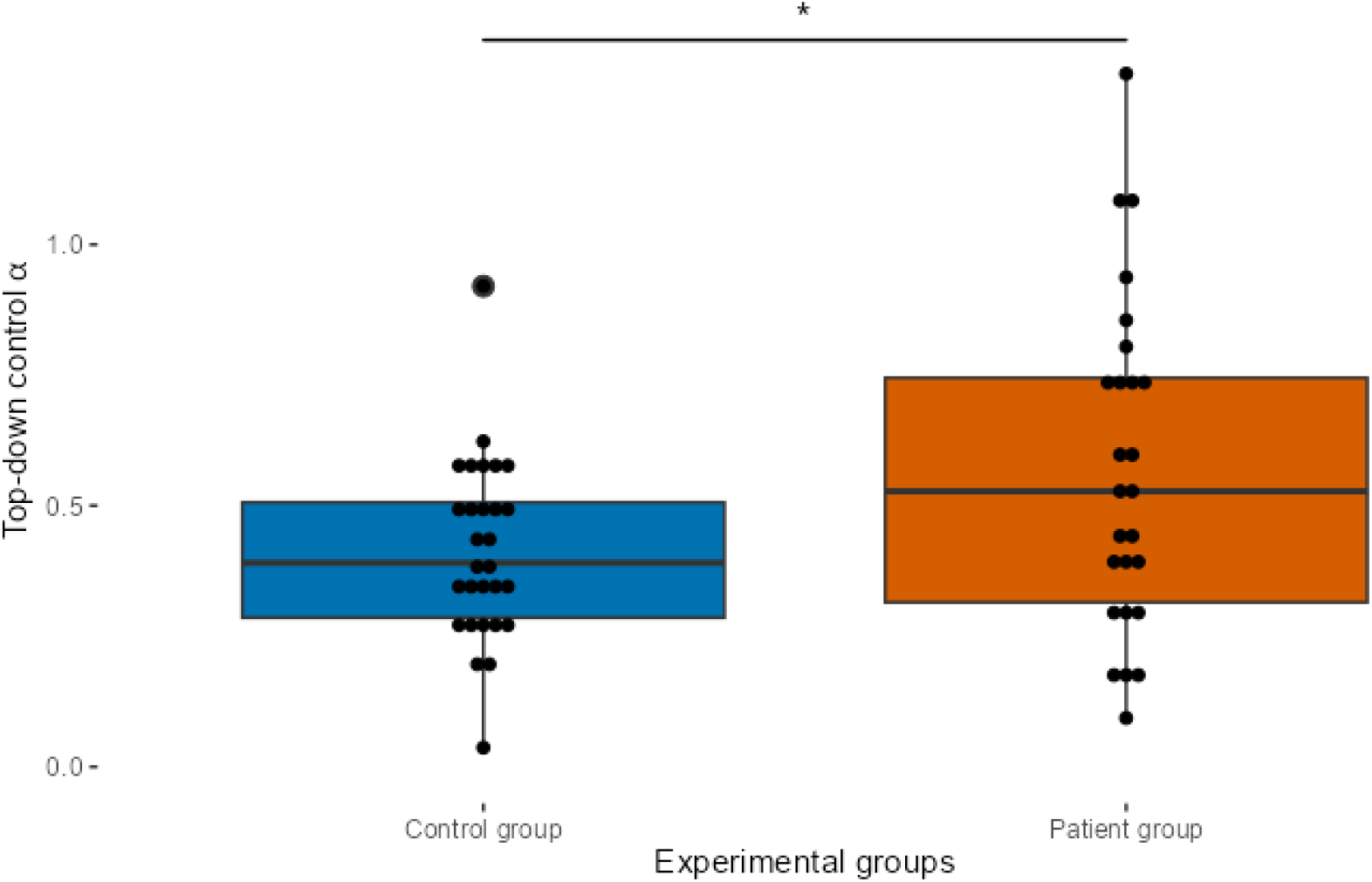
Group differences in top-down control. α. aMCI patients (orange) showed worse top-down control α than control participants (blue; *t*(37.43) = -2.24, *p* = .031). The parameter α represents selectivity (inverted scale). Thus, values closer to 1 represent *worse* top-down control and values closer to 0 *better* top-down control. Each dot in the boxplot depicts the data of a single study participant. The horizontal line in each box represents the median value per group. The larger dot above the control group boxplot signifies an outlier data point exceeding 1.5 interquartile range. **Figure 2. Alt text.** Boxplot on the group difference of top-down control alpha between the control group and the patient group. The average of the control group is significantly lower, which is marked with an asterisk above the boxplot.

**Table 2.** Key behavioural and functional connectivity measures by group.

|  | **Control group** | **aMCI Patient group** | **Group differences** |
|---|---|---|---|
|  | n = 29 | n = 26 |  |
| **Behavioural and functional connectivity measures** |  |  |  |
| Top-down control α, mean (SD) [lower values indicate better control] | 0.41 (0.17) | 0.57 (0.32) | *t*(53) = -2.24\* |
| *l*FPN-rsFC cluster **z**-value, mean (SD) | 0.55 (0.87) | -0.61 (0.76) | *t*(53) = 5.28\*\*\* |
| VN-rsFC cluster **z**-value, mean (SD) | 0.38 (1.16) | -0.42 (0.56) | *t*(41.29) = 3.31\*\* |
| *r*FPN-rsFC cluster **z**-value, mean (SD) | 0.35 (1.05) | -0.39 (0.8) | *t*(53) = 2.94\*\* |

*l*FPN-rsFC, VN-rsFC, and *r*FPN-rsFC refer to the average functional connectivity as **z**-values within the cluster in the left frontoparietal network, the visual network, and the right frontoparietal network, respectively.
aMCI: amnestic mild cognitive impairment.
\**P* < 0.05, \*\**P* < 0.01, \*\*\**P* < 0.001.

### Network selection and cluster analysis

The dual regression and subsequent Allen et al. template (51) correlation resulted in 18 components identified as resting-state networks (see Additional file 4). Initially, six of those components were labelled as attention networks and four separate components as visual networks (Table 3). The results of the NCT led to dropping two attention networks, as they corresponded to a sensorimotor and a language network. The remaining four attention networks and the four visual networks identified with the Allen et al. templates (51) were confirmed with the NCT. The final four attention networks selected were labelled as left and right frontoparietal networks (IC_00 and IC_04, respectively), the cingulo-opercular network (IC_12), and the dorsal attention network (IC_15). Visual networks (IC_03, IC_06, IC_14, IC_30) were labelled as visual networks.

**Table 3.** Attention and visual networks.

| **Network name in Allen** (51) | **ICA component in our data** | **Correlation with Allen** (51) | **Correspondence with Yeo** (17, 71) **according to NCT** (52) **(dice coefficient)** |
|---|---|---|---|
| **Attention networks** |  |  |  |
| Left FPN | IC-00 | *r* = 0.33 | Control B (0.22)** |
| Right FPN | IC-04 | *r* = 0.36 | Control B (0.29)** |
| CON | IC-12 | *r* = 0.35 | Salience / Ventral Attention B (0.33)** |
| Precuneus network | IC-15 | *r* = 0.30 | Dorsal Attention A (0.16)* |
| *Precuneus network* | *IC-17* | *r = 0.30* | *Somatomotor A (0.41)*\*\* |
| *TPJ network, alerting system* | *IC-28* | *r = 0.30* | *Temporoparietal (0.13)*\*\* |
| **Visual Networks** |  |  |  |
| VN 1 | IC-03 | *r* = 0.36, *r* = 0.32<sup>a</sup> | Visual A (0.33)*** |
| VN 2 | IC-06 | *r* = 0.47, *r* = 0.41<sup>a</sup> | Visual B (0.33)** |
| VN 3 | IC-14 | *r* = 0.32 | Visual A (0.33)* |
| VN 4 | IC-30 | *r* = 0.42 | Visual A (0.22) |

Networks presented in italics were discarded after using NCT due to better correspondence with other networks.
ICA: independent component analysis, NCT: network correspondence toolbox, FPN: frontoparietal network, IC: independent component, CON: cingulo-opercular network, TPJ: temporo-parietal junction, VN: visual network.
<sup>a</sup>VN 1 and VN 2 correspond with two visual networks defined by Allen.
\**P* < 0.05, \*\**P* < 0.01, \*\*\**P* < 0.001.

Regarding the cluster analysis, two clusters emerged, where controls displayed significantly higher rsFC values than patients (Fig. 3, Table 2). The first cluster (Fig. 3A; *p* = 0.001) was found in the left frontoparietal network (*l*FPN, IC_00), overlapping with the inferior parietal lobule (IPL), specifically the supramarginal gyrus. The second cluster (Fig. 3B; adjusted *p* = 0.016) was found in one of the visual networks (VN, IC_03) and overlapped with the bilateral ventral occipital cortex. Initially, after applying TFCE, four clusters were also found in the visual network but only one survived correction for multiple comparisons (false discovery rate of 0.05).

**Figure 3.**
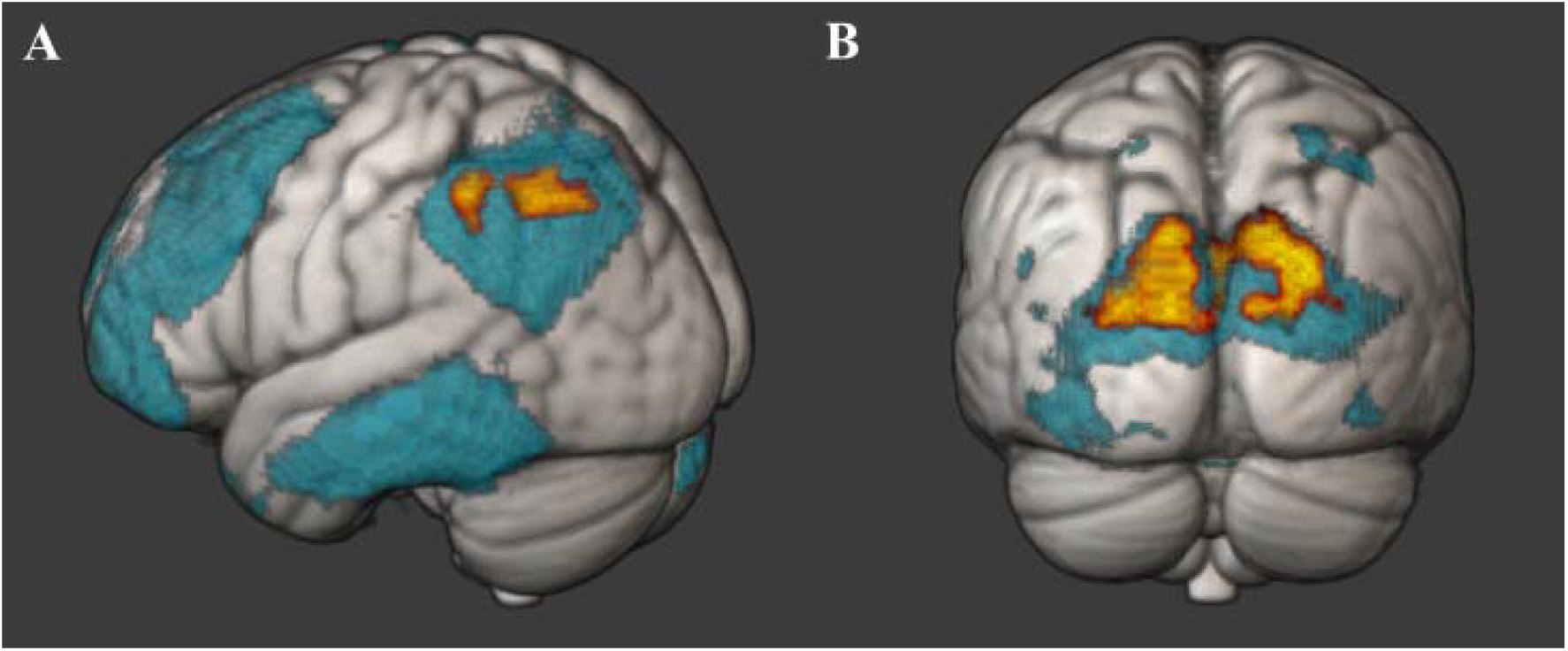
Clusters in resting-state networks where group differences in functional connectivity (rsFC) were found. Patients with aMCI showed significantly lower rsFC than healthy control participants in two clusters (yellow) in two networks (blue). (**A**) The first cluster (*k* = 243 vx, p = 0.001) was found within the inferior parietal lobule of the left frontoparietal network. (**B**) The second cluster (k = 2342 vx, p = 0.004) was found within the bilateral occipital lobe of a visual network. **Figure 3. Alt text.** Figure labelled A and B and depicting a 3D brain in each panel. The brains are used to display a brain network each and one cluster in the network where significant differences between groups were found. A depicts a brain from the left side and showing a bit of the occipital lobe. Parts of the left temporal lobe, frontal lobe and parietal lobe are marked blue. Two small parts of the parietal lobe are marked yellow. B depicts a brain from behind. Parts of the bilateral occipital lobe are marked blue. A large bilateral, dorsal cluster within the blue part is marked yellow.

### Correlation between rsFC in attention and visual networks and top-down control α

We found a significant negative correlation between rsFC in *l*FPN and top-down control α across all participants (*ρ* = -0.33, *p* = 0.014, 95 % CI [-0.59, -0.02], Fig. 4A) and in the patient group (*ρ* = -0.48, *p* = 0.014, 95 % CI [-0.74, -0.1], Fig. 4B), indicating that lower *l*FPN-rsFC is associated with worse top-down control α. No significant correlation was found for the control group (*ρ* = -0.08, *p* = 0.914, 95 % CI [-0.44, 0.31], Fig. 4B). When Group was controlled for, no significant correlation was found between *l*FPN-rsFC and top-down control α (*ρ* = -0.237, *p* = 0.084), suggesting that Group might influence the relationship between *l*FPN-rsFC and top-down control α. In the visual network (VN), rsFC did not significantly correlate with top-down control α (*ρ_total_* = -0.18, *p* = 0.185, 95 % CI [-0.42, 0.08], Fig. 4C, *ρ_patients_* = -0.03, *p* = 0.898, 95 % CI [-0.42, 0.38], *ρ_controls_* = -0.02, *p* = 0.914, 95 % CI [-0.39, 0.36], Fig. 4D).

**Figure 4.**
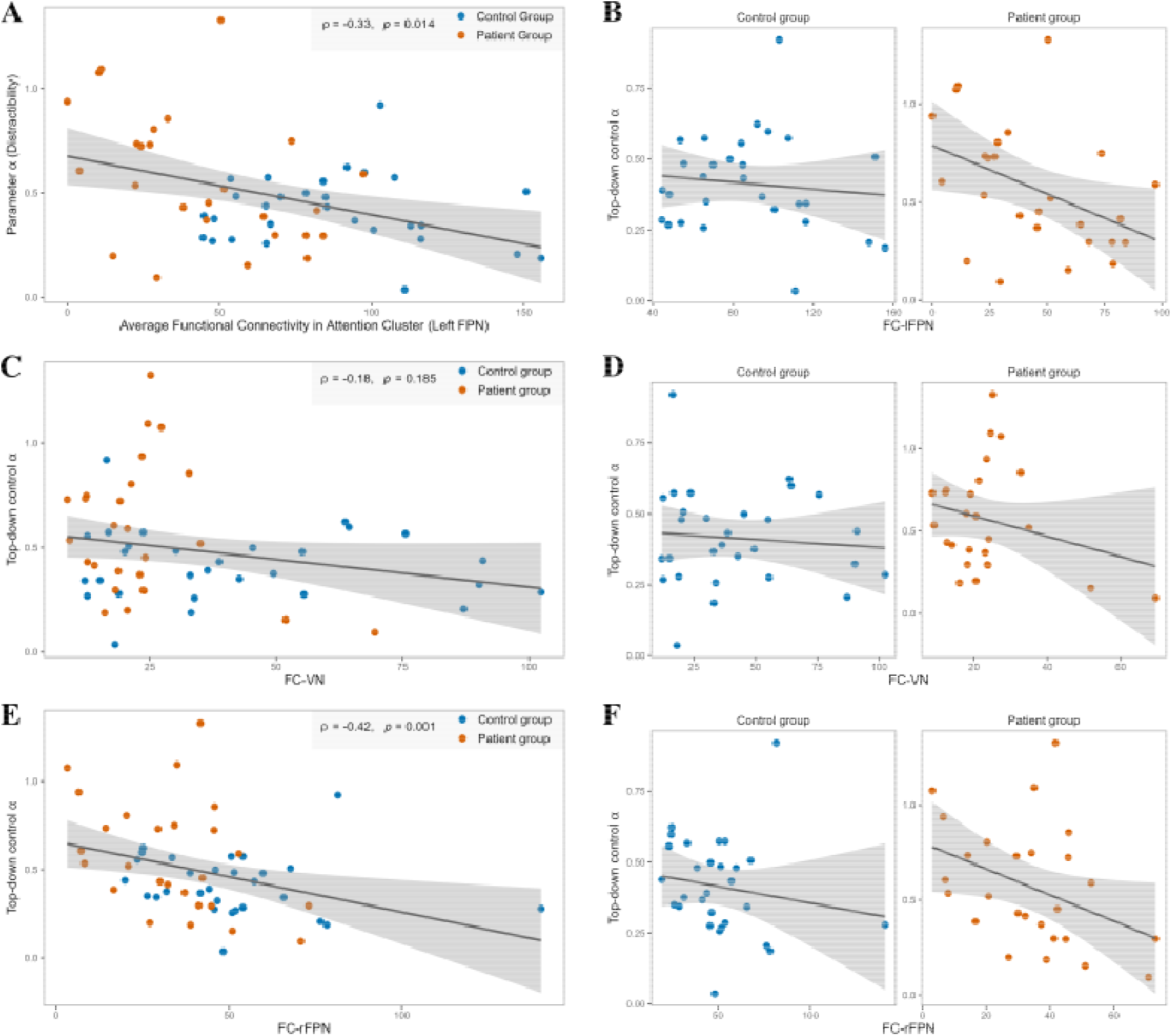
Association of functional brain connectivity (rsFC) with top-down control α. (**A**) Significant negative correlation between *l*FPN-rsFC and top-down control α across participants (*ρ* = -0.33, *p* = 0.014) (**B**) and in the aMCI patient group (*ρ* = -0.48, p = 0.014). (**C**) No significant correlation between VN-rsFC and top-down control *α* emerged, neither across participants (*r* = -0.18, *p* = 0.185) (**D**) nor in any group (ρ_controls_ = -0.02, p = 0.914; *ρ*_patients_ = -0.03, p = 0.898). (**E**) Significant negative correlation between *r*FPN-rsFC and top-down control α across participants (*r* = -0.42, *p* = 0.001) (**F**) and in the aMCI patient group (*ρ* = -0.40, p = 0.042). X-axis: z-values representing rsFC; note the differing axis limits. Y-axis: higher values indicate worse top-down control α. **Figure 4. Alt text.** Correlation figures labelled A to F. Panels A, C and E depict scatterplots of the entire sample with a regression line and standard error margins. Functional connectivity of the left frontoparietal network, the visual network and the right frontoparietal network is correlated with top-down control alpha. In the scatterplot, patients and controls are assigned different colours. The correlation coefficient r and the p-value is written in the top right corner of each panel. Panels B, D and F depict one scatterplot per group each with a regression line and standard error margins. Functional connectivity of the left frontoparietal network, the visual network and the right frontoparietal network is correlated with top-down control alpha.

### Mediation of top-down control

We tested whether *l*FPN-rsFC mediates the association between group and top-down control α (Fig. 5). The indirect effect (ab: 0.1, SE = 0.05, 95% CI [0.01, 0.21) was significant, indicating that group differences in top-down control α were mediated by group differences in *l*FPN-rsFC.

**Figure 5.**
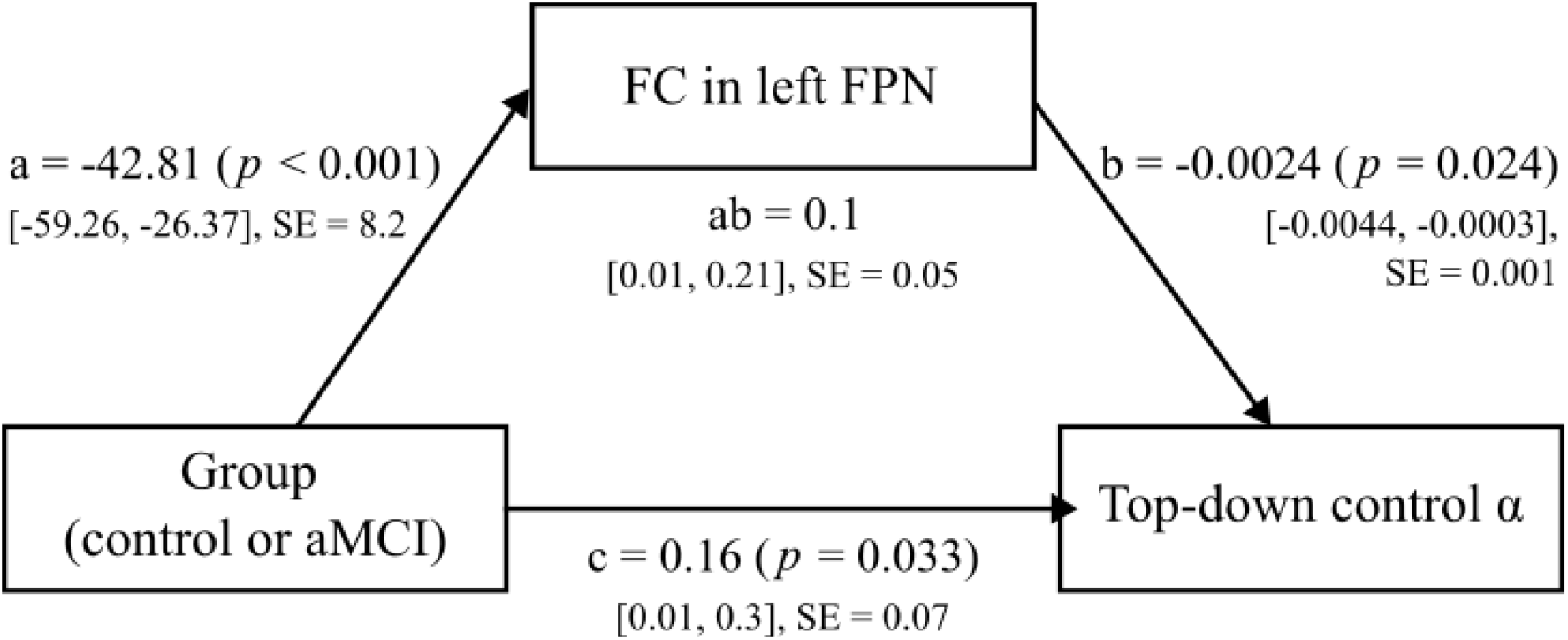
Mediation model of group differences in top-down control. α. Group was entered as the predictor, functional connectivity in left FPN (*I*FPN-rsFC) as the mediator, and top-down control α as the outcome. The indirect effect of group on top-down control α through *l*FPN-rsFC was significant. All paths are described by their effect estimates, significance values in parentheses, 95 % confidence intervals in brackets and the standard errors (SE). **Figure 5. Alt text.** Schematic of the mediation analysis. In the bottom left corner is a box with the label ‘Group (control or aMCI)’, from which two arrows point towards two other boxes. One box is in the bottom right corner with the label ‘Top-down control alpha’ and another box is at the top centre with the label ‘rsFC in left FPN’. Another arrow points from the top centre box towards the bottom right box. Each arrow has path information from the analysis written next to it. Additional model information is below the top centre box.

### Control analyses

The partial correlations using age, years of education and depressive symptoms as control variables revealed results consistent with the bivariate analyses. More specifically, *l*FPN-rsFC and top-down control α correlated significantly across participants (ρ = -0.34, *p* = 0.013) and in the patient group (*ρ* = -0.46, *p* = 0.027), but not in the control group (*ρ* = -0.17, *p* = 0.429). Likewise, VN-rsFC did not significantly correlate with top-down control *α* in a partial correlation, neither across participants (*ρ* = -0.18, *p* = 0.209), nor in the patient (*ρ* = 0.01, *p* = 0.956) or control group (*ρ* = 0.00, *p* = 0.997).

As significant patient-control differences in rsFC emerged only in the *l*FPN, we tested whether the relationship with top-down control α showed hemisphere-specificity. We mirrored the *l*FPN cluster’s coordinates using *fslswapdim* to create a mirrored cluster in the right frontoparietal network (*r*FPN, IC_04). This cluster is located in the inferior parietal lobule but corresponds more closely to the angular gyrus according to the Harvard-Oxford Cortical Structural Atlas in *fsleyes*. We then correlated the rsFC values of the *r*FPN (*r*FPN-rsFC) with those of top-down control α. We found a significant negative correlation of *r*FPN-rsFC and top-down control α for the total sample (*ρ* = -0.42, *p* = 0.001, 95 % CI [-0.63, -0.18], Fig. 4E) and the patient group (*ρ* = -0.40, *p* = 0.042, 95 % CI [-0.69, -0.004], Fig. 4F) but not the control group (*ρ* = -0.28, *p* = 0.139, 95 % CI [-0.6, 0.11], Fig. 4F). These results indicate that the relationship between top-down control *α* and rsFC in the frontoparietal attention network is not specific to the left hemisphere. The correlation between *r*FPN-rsFC and top-down control *α* remained significant when group was used as a control variable (*ρ* = - 0.36, *p* = 0.007), suggesting a more general relationship between *r*FPN-rsFC and top-down control α.

*r*FPN-rsFC and top-down control α correlated significantly across participants when age, education and depressive symptoms were controlled for (*ρ* = -0.42, *p* = 0.002), although no significant correlation was found in the patient group (*ρ* = -0.36, *p* = 0.088) or control group (*ρ* = -0.38, *p* = 0.058), separately.

## Discussion

In the present study, we investigated the association between resting state functional brain connectivity and top-down attentional control in a sample of patients with aMCI and healthy control participants. We found both worse top-down control α in patients with aMCI and reduced rsFC in one attention (*l*FPN) and one visual network. Lower *l*FPN-rsFC was associated with worse top-down control α across participants and within the patient group, and this association remained significant after controlling for age, years of education and depressive symptoms. An exploratory analysis of the homologous right-hemisphere area revealed a comparable association with top-down control. In addition, an exploratory mediation analysis indicated that *l*FPN-rsFC statistically mediated group differences in top-down control α. In contrast, VN-rsFC was not associated with top-down control, suggesting a more specific relationship between frontoparietal network integrity and impaired attentional selection in aMCI. Together, these findings extend the clinical characterization of aMCI beyond its defining memory impairment by identifying a specific deficit in goal-directed attentional selection and linking it to altered frontoparietal network function.

Worse top-down control α in patients with aMCI indicates a stronger bias of attentional selection towards distractor stimuli (8,56,57). Across all participants and within the patient group, worse top-down control was associated with reduced rsFC in the *l*FPN, and this association remained significant after controlling for age, education and depressive symptoms. The primary network analysis identified a significant patient-control difference only within the *l*FPN, while no corresponding cluster emerged in the *r*FPN. In an exploratory control analysis, however, rsFC in the homologous *r*FPN was also associated with top-down control α across participants and in the patient group. This exploratory finding suggests that the association between frontoparietal connectivity and attentional top-down control may not be strictly left-lateralized. Previous evidence has linked activity (21,58,59) and connectivity (60) in frontoparietal areas with performance in visual attention tasks in healthy older adults and older adults at risk of AD dementia. Our results extend these findings by demonstrating an association between rsFC within frontoparietal areas and a process-specific component of visual attention function, top-down control α, in clinically defined aMCI. This is clinically relevant because aMCI is commonly characterized primarily by memory impairment, although patients may also experience deficits in attentional and executive control that are not fully captured by memory-centred clinical characterization. The present findings suggest that process-specific measures of attentional selection may provide complementary information about cognitive dysfunction in aMCI. More broadly, our findings are consistent with the proposed role of frontoparietal control hubs in cognitive reserve. The modal control hub framework suggests that these hubs may support cognitive performance by facilitating transitions to cognitively demanding brain states (61). The association between FPN-rsFC and top-down attentional control observed here provides a behavioural correlate of FPN integrity that is compatible with this framework, although cognitive reserve itself was not directly assessed. This broader interpretation is also consistent with previous studies linking FPN integrity to cognitive decline in Alzheimer’s disease (60,62,63) and normal aging (59,64).

Beyond the associations, an exploratory mediation analysis indicated that *l*FPN-rsFC statistically mediated group differences in top-down control α. Altered *l*FPN connectivity has previously been reported in the MCI stage (22,65). Together with the observed association between *l*FPN and top-down control α, the mediation model is consistent with the possibility that reduced frontoparietal network connectivity contributes to impaired top-down attentional control in aMCI. Given the cross-sectional design, however, a causal contribution of altered frontoparietal connectivity to impaired top-down control cannot be established. Within the neural interpretation of TVA, nTVA, top-down filtering is thought to bias competition between objects by modulating the neural resources allocated to object representations in favour of task-relevant objects (66). Reduced filtering efficiency may therefore increase the likelihood that task-irrelevant information gains access to visual short-term memory.

Patients with aMCI also showed reduced rsFC in the VN. Although hypoactivation and reduced rsFC in visual areas have been shown in AD, evidence for such changes at the MCI stage is more limited (18,67). Importantly, no association of reduced rsFC in the VN with top-down control α was found, suggesting that the relationship with attentional selectivity was more specifically related to FPN connectivity. This dissociation is important because it indicates that reduced connectivity in aMCI was not uniformly related to attentional performance: despite group differences in both networks, only frontoparietal connectivity was associated with the process-specific attentional deficit.

Nevertheless, the concurrent reductions of rsFC in the *l*FPN and the VN may reflect broader changes in interacting systems involved in attentional selection. Frontoparietal control signals are known to modulate the processing of visual input (68), and the neural interpretation of TVA assigns the pulvinar nucleus in the thalamus a central role in representing attentional priority and regulation competition between objects (66). Given the strong functional interactions of the pulvinar nucleus to parietal and visual occipital regions (69), future studies could investigate whether alterations in these systems are functionally coupled in aMCI. More broadly, reduced rsFC across both frontoparietal and visual networks is consistent with previous evidence of distributed functional connectivity alterations in MCI (70).

From a clinical perspective, the present findings emphasize that cognitive dysfunction in aMCI may extend beyond episodic memory. The absence of significant associations between top-down control α and global cognitive scores or hippocampal volume further suggests that the attentional deficit captured by α may reflect information that is not redundant with conventional measures of global cognition or structural medial temporal lobe change. Process-specific assessment may therefore help characterize cognitive heterogeneity within clinically defined aMCI, although its diagnostic or prognostic utility remains to be established in larger longitudinal cohorts.

Our findings should be interpreted taking some limitations into consideration. Firstly, the cross-sectional nature of our study precludes us from making causal inferences regarding the association of *l*FPN and top-down control and our relatively small sample limited statistical power, particularly for subgroup and interaction analyses. Moreover, aMCI was clinically defined and biomarker information was available only for a subset of patients. The present findings should therefore be interpreted as correlates of clinical aMCI rather than as specific markers of underlying AD pathology. This also means that the sample likely represents etiologically heterogeneous aMCI, which is relevant to the generalizability of the findings but precludes conclusions about AD-specific network mechanisms.

Secondly, the anatomical T1-weighted acquisition protocol was updated during patient recruitment, whereas functional image acquisition remained unchanged. Sensitivity analyses indicated that the main *l*FPN group difference remained significant when analyses were restricted to patients scanned with the same anatomical protocol as the control group; nevertheless, an influence of the protocol change on preprocessing and spatial normalization cannot be entirely excluded. Finally, our sample included more men than women, precluding a systematic examination of potential gender differences in the relationship between rsFC and attentional control. Likewise, the relatively high educational level of our sample may limit the generalisability to more socioeconomic and ethnically diverse populations.

## Conclusion

We found both worse top-down control α and reduced rsFC in one frontoparietal and one visual network in patients with aMCI compared with cognitively unimpaired older adults. Reduced rsFC in the *l*FPN was associated with poorer top-down control and statistically mediated the group difference in α. Our results identify reduced frontoparietal rsFC as a neural correlate of impaired attentional selectivity in aMCI and further indicate that cognitive alterations at this stage extend beyond memory to goal-directed attentional control. More broadly, these findings expand upon previous evidence for a role of the FPN in supporting cognitive control and maintaining cognitive performance.

## Declarations

### Data availability

The behavioural dataset as well as the fMRI dual regression stage 2 and 3 data generated and analysed during the current study are available in the ZENODO repository, 10.5281/zenodo.23801723. Other fMRI data generated and analysed during the current study are available in the NEUROVAULT repository, https://neurovault.org/collections/24650/.

## Funding

This project has received funding from the IZKF Advanced Medical Scientist Program of Jena University Hospital and Fit4BRAIN has received funding from the European Union’s Horizon 2020 research and innovation program under the Marie Skłodowska-Curie grant agreement no 859890.

## Competing interests

The authors report no competing interests.

## Author’s contributions

H.K.: Conceptualization, Formal analysis, Investigation, Data curation, Visualization, Writing – original draft. K.F.: Conceptualization, Funding acquisition, Project administration, Supervision, Writing – original draft. F.K.: Investigation, Data curation, Writing – review & editing. M.H.: Investigation, Writing - review & editing. S.J.S.: Investigation, Data curation, Writing – review & editing. S.B.: Resources, Writing - review & editing. H.-Y.C.: Resources, Writing - review & editing. A.R.-R.: Conceptualization, Funding acquisition, Project administration, Supervision, Formal analysis, Writing – original draft.

## Supplementary material

Supplementary material is available. This includes additional analyses and 3 supplementary tables.

## List of Abbreviations

ACE-III: Addenbrooke’s Cognitive Examination III
AD: Alzheimer’s disease
aMCI: amnestic Mild Cognitive Impairment
BADL: Bayer Activities of Daily Living Scale
BADS-DEX: Behavioural Assessment of the Dysexecutive Syndrome Questionnaire
BBR: Boundary-Based Registration
BOLD: blood level oxygen dependent
CDR: Clinical Dementia Rating
CERAD: Consortium to Establish a Registry for Alzheimer’s Disease
CI: confidence interval
CSF: cerebrospinal fluid
DMN: default mode network
EPI: echo planar imaging
fMRI: functional magnetic resonance imaging
FWIT: Stroop task
GDS: Geriatric Depression Scale
HADS-D: Hospital Anxiety and Depression Scale
IC: independent component
ICA: independent component analysis
IPL: inferior parietal lobule
IQCODE: Information Questionnaire on Cognitive Decline for the Elderly
*l*FPN: *left* frontoparietal network
MCI: Mild Cognitive Impairment
MNI: Montreal Neurological Institute
MP-RAGE: magnetization-prepared rapid gradient echo
MRI: magnetic resonance imaging
MWT-B: “Mehrfachwahl-Wortschatz-Intelligenztest“ version B
NCT: Network Correspondence Toolbox
nTVA: neural interpretation of the Theory of Visual Attention
PSQI: Pittsburgh Sleep Quality Index
*r*FPN: *right* frontoparietal network
ROCFT: Rey Osterrieth Complex Figure Test
rsFC: resting-state functional connectivity
SE: standard error
TFCE: threshold-free cluster enhancement
TMT: Trail-Making-Test
TVA: Theory of Visual Attention
VN: visual network
ZST: Digit Symbol Substitution Test

## Notes

### Competing Interest Statement

The authors have declared no competing interest.

### Author Declarations

The Ethics Committee of Jena University Hospital gave ethical approval for this work.

