## Supplementary Material for "Frontoparietal functional connectivity underlies top-down attention in mild cognitive impairment"

### Supplementary Analyses

#### AD biomarkers in CSF and MRI

Out of 26 patients, CSF biomarker data were available for 20 (76.9 %) patients with aMCI. Six patients showed clear AD-related pathology (AD+, decreased Amyloid β and either standard or elevated tau levels) and eight patients showed no AD-related pathology (AD-). The results of four patients were categorized as ‘unclear’ due to showing only elevated tau levels and excluded from analysis. Clinical MRI data were available for 23 (88.5%) patients with aMCI. Fifteen patients showed MRI abnormalities related to neurodegeneration (MRI+, general atrophy, enlarged ventricles) and 8 patients showed no such abnormalities (MRI-). Neither top-down control *α* nor rsFC differed between CSF or MRI groups (all *p*-values ≥ 0.137). Combining CSF and MRI data, we separated patients into groups showing either pathology (AD+ or MRI+ or both) or no pathology (AD- and MRI-). Again, no differences in top-down control *α* or rsFC were found (all *p*-values ≥ 0.200).

#### Sensitivity analyses for MRI sequence update

We estimated mean rsFC within the entire *l*FPN (IC-00) and VN (IC-03) and found no significant differences between patients measured before and after the anatomical sequence update (*l*FPN: *t*(24) = 0.53, *p* = 0.605; VN: *t*(24) = -0.68, *p* = 0.511). Within the clusters showing patient-control differences, patients measured before the update had significantly lower rsFC values within the *l*FPN cluster (*t*(24) = -2.34, *p* = 0.028) whereas no significant difference was found in the VN cluster (*t*(24) = -1.08, *p* = 0.300). To test whether the primary patient-control differences in rsFC were dependent on the anatomical acquisition sequence, we repeated the comparisons including only participants measured with the updated sequence, which was also used in controls. The differences in the *l*FPN-rsFC cluster remained significant (12 patients vs. all 29 controls; *t*(39) = -2.88, *p* = 0.006). The VN-rsFC difference was no longer significant (*t*(39) = -1.81, *p* = 0.078) although mean rsFC remained numerically lower in patients (mean patients: 26.46, mean controls: 41.88).

### Supplementary Tables

**Supplementary Table 1 Distribution of AD biomarkers and MRI neurodegeneration profiles in patients with aMCI**

|  | **Atrophy according to MRI (n = 23)** | |
| --- | --- | --- |
| **CSF AD biomarkers** | **Present (MRI+, n =15)** | **Absent (MRI-, n= 8)** |
| Aβ and tau present (Aβ+T+) | 4 | 0 |
| Aβ present and tau absent (Aβ+T-) | 2 | 0 |
| Aβ absent and tau present (Aβ-T+) | 4 | 0 |
| Aβ and tau absent (Aβ-T-) | 3 | 7 |
| No CSF data available | 2 | 1 |

Biomarker CSF and MRI data were obtained from medical records. Three patients refused both MRI and the lumbar puncture procedure and have been excluded from this overview. According to medical records, CSF biomarker data indicates AD when Aβ levels are decreased and tau levels are elevated (described here as Aβ+T+). Decreased Aβ and standard tau levels generally indicate an earlier AD stage (Aβ+T-). Standard Aβ levels generally indicate an absence of AD biomarkers but are classified as ‘unclear’ pathology in combination with elevated tau levels (Aβ-T+). Standard Aβ and standard tau levels indicate an absence of AD biomarkers (Aβ-T-).

AD: Alzheimer’s disease, MRI: magnetic resonance imaging, aMCI: amnestic mild cognitive impairment, CSF: cerebrospinal fluid, Aβ: Amyloid beta

**Supplementary Table 2 Association of top-down control *α* with demographic variables, neuropsychological test results, neuropsychiatric symptoms, and hippocampal volume**

|  | **Total** | **Control group** | **aMCI Patient group** |
| --- | --- | --- | --- |
|  | *N* = 55 | *n* = 29 | *n* = 26 |
| **Demographic variables** | | |  |
| Age | *ρ* = 0.002 | *ρ* = 0.03 | *ρ* = -0.08 |
| Education | *ρ* = -0.20 | *ρ* = -0.20 | *ρ* = -0.16 |
| **Neuropsychological test results** | | | |
| ACE-III | *ρ* = -0.10 | *ρ* = -0.22 | *ρ* = 0.35 |
| CERAD Chandler | *ρ* = -0.10 | *ρ* = -0.04 | *ρ* = 0.13 |
| **Neuropsychiatric symptoms** | | | |
| HADS Anxiety | *ρ* = 0.09 | *ρ* = -0.14 | *ρ* = 0.14 |
| HADS Depression | *ρ* = 0.08 | *ρ* = -0.27 | *ρ* = 0.15 |
| **Hippocampal volume** | | | |
| Left hippocampal volume (mm³) | *ρ* = -0.08 | *ρ* = -0.03 | *ρ* = 0.04 |
| Right hippocampal volume (mm³) | *ρ* = -0.09 | *ρ* = -0.01 | *ρ* = 0.02 |

ACE-III: Addenbrookes Cognitive Examination III, CERAD: Consortium to Establish a Registry for Alzheimer’s Disease, HADS: Hospital Anxiety and Depression Scale.

**P* < 0.05, ***P* < 0.01, ****P* < 0.001.

**Supplementary Table 3 All resting-state networks**

| **ICA component in our data** | **Network according to Allen** | **Correlation with Allen** | **Network image** |
| --- | --- | --- | --- |
| **IC-00** | Attention network, *l*FPN | *r* = 0.33 | 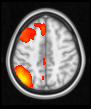 |
| **IC-01** | Sensorimotor network | *r* = 0.34 | 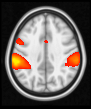 |
| **IC-02** | DMN | *r* = 0.35 | 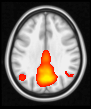 |
| **IC-03** | Visual network | *r* = 0.36 | 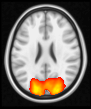 |
| **IC-04** | Attention network, *r*FPN | *r* = 0.36 | 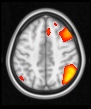 |
| **IC-05** | DMN | *r* = 0.36 | 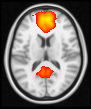 |
| **IC-06** | Visual network | *r* = 0.47 | 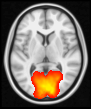 |
| **IC-07** | Sensorimotor network | *r* = 0.55 | 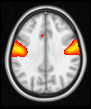 |
| **IC-09** | Auditory network | *r* = 0.33 | 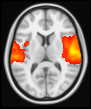 |
| **IC-12** | Attention network, CON | *r* = 0.35 | 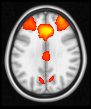 |
| **IC-12** | DMN | *r* = 0.30 |  |
| **IC-13** | Sensorimotor network | *r* = 0.40 | 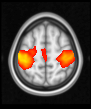 |
| **IC-14** | Visual network | *r* = 0.32 | 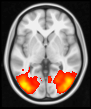 |
| **IC-15** | Attention network | *r* = 0.30 | 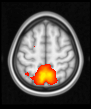 |
| **IC-17** | Sensorimotor network | *r* = 0.47 | 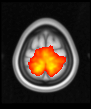 |
| **IC-17** | Attention network | *r* = 0.30 |  |
| **IC-24** | Sensorimotor network | *r* = 0.42 | 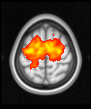 |
| **IC-28** | Attention network | *r* = 0.30 | 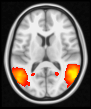 |
| **IC-30** | Visual network | *r* = 0.42 | 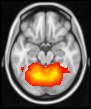 |
| **IC-43** | Basal ganglia | *r* = 0.37 | 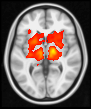 |
| **IC-65** | DMN | *r* = 0.30 | 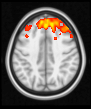 |

ICA: independent component analysis, *l*FPN: left frontoparietal network, DMN: default mode network, *r*FPN: right frontoparietal network, CON: cingulo-opercular network.
